# Japanese version of the Family Stigma Instrument for informal caregivers of people with dementia

**DOI:** 10.64898/2026.03.12.26348133

**Authors:** Taiji Noguchi, Jem Bhatt, Ayane Komatsu, Ryota Watanabe

**Author notes:** Corresponding author: Taiji Noguchi, PhD, Department of Community Health and Preventive Medicine, Hamamatsu University School of Medicine 1-20-1 Handayama, Chuoku, Hamamatsu, Shizuoka 431-3192.

## Abstract

**INTRODUCTION:** Overcoming dementia-related stigma is a global challenge, but tools to assess stigma among family caregivers of people living with dementia remain limited. This study examined the validity and reliability of the Japanese version of the Family Stigma Instrument for family caregivers of people living with dementia (J-FAMSI-dementia), originally developed in the United Kingdom.

**METHODS:** A total of 372 informal caregivers aged 18–79 years of family members living with dementia completed an internet survey. The J-FAMSI-dementia comprises five subscales (stigma by association; perceived, affective, and behavioral affiliate stigma; and positive aspects of caregiving), developed through forward and back translations.

**RESULTS:** Confirmatory factor analysis supported an acceptable five-factor model. All subscales showed high internal consistency and moderate to good test-retest reliability. Correlations with dementia attitude, caregiving burden, and depressive symptoms supported construct validity.

**DISCUSSION:** The J-FAMSI-dementia demonstrated acceptable validity and reliability and may help identify dementia-related stigma among family caregivers.

**Highlights:**

- The Family Stigma Instrument for dementia in Japan (J-FAMSI-dementia) was developed
- The J-FAMSI-dementia showed acceptable validity and good reliability
- The scale included associative and affiliate stigma and positive caregiving aspects
- The J-FAMSI-dementia helps understand the dementia stigma experienced by caregivers

**Research in context:**

1. **Systematic review:** Despite reducing dementia-related stigma being an international priority, previous research has mainly focused on public stigma and self-stigma, with less attention to stigma experienced by family caregivers.
2. **Interpretation:** This study developed a Japanese version of the Family Stigma Instrument for dementia (J-FAMSI-dementia), originally developed in the United Kingdom, and provided a tool to assess stigma experienced by informal caregivers of people living with dementia in Japan.
3. **Future directions:** The J-FAMSI-dementia, with demonstrated validity and reliability, may help in understanding the stigma experienced by caregivers in dementia care and facilitate efforts to reduce stigma and strengthen caregiver support.

## BACKGROUND

Dementia is a degenerative neurological disorder that affects an estimated 57.4 million people worldwide in 2019 and is projected to affect 152.8 million by 2050.^1^ This rapid increase not only impacts the individuals living with dementia and health care systems, but also places enormous burdens on their informal caregivers.^2,3^ Japan, a fast-aging country, has one of the highest rates of dementia in the world. Nearly seven million Japanese people were living with dementia in 2025,^4^ with projections suggesting that one-third of the population will be affected by dementia by 2060.^5^ In response to these public health issues, many countries have developed policies to support people living with dementia. For instance, the new Dementia Strategy for Scotland aims to create a society centered on people living with dementia and their care partners, emphasizing overall societal action as well as health and social care.^6^ In Japan, the Basic Act on Dementia to Promote an Inclusive Society, enacted in 2023, seeks to comprehensively enable people living with dementia and their families to live with dignity and lead meaningful lives.^7^ These share the goal of promoting inclusive societies for both people living with dementia and their families.

Addressing the stigma of dementia is a key challenge to achieving such an inclusive society. Stigma has been conceptualized as a mark of social disgrace whereby the targeted individuals are discredited because of attributes such as ethnicity, mental health problems, illness, disability, or drug use.^8^ It manifests as prejudice and discrimination. Globally, approximately 80% of people living with dementia experience some form of discrimination, an increase in recent years.^9^ Because stigma leads to a decline in quality of life of both the individuals and their families and is a crucial obstacle that restricts dignity and human rights, overcoming dementia-related stigma is an international issue.

The framework of four forms of stigma is well-accepted.^10^ First, public stigma refers to the attitudes held by society toward stigmatized individuals and groups.^11,12^ Second, self-stigma occurs when stigmatized persons recognize and internalize the public stigma.^12,13^ Third, institutional stigma arises when policies restrict the choices and opportunities of stigmatized individuals.^14,15^ The fourth type is referred to as family stigma, also called stigma by association or courtesy stigma, in which people associated with the target individual are stigmatized.^16,17^ The persons associated with the stigmatized person, particularly family caregivers, may be blamed for the person’s illness or disability and may themselves experience prejudice and discrimination.^8^ Furthermore, the process of persons associated with the stigmatized individual internalizing stigma is referred to as affiliate stigma^17,18^;it comprises three interrelated psychological responses, including perceptions, affect, and behavior aspects.^19^ Research has primarily focused on self-stigma among stigmatized individuals and public stigma in the general population. However, the stigma among family caregivers of people living with dementia has often been overlooked. Family stigma can lead to negative feelings and perceptions of family members about themselves, resulting in social withdrawal and concealment.^9,17,18^ Therefore, focusing on the stigma among family caregivers, understanding its realities, and exploring appropriate support strategies are required.

Although several stigma scales for family caregivers of people living with dementia have been developed,^19,20^ assessment scales that comprehensively capture family stigma experiences remain limited. Chang et al. standardized the Affiliate Stigma Scale to assess affiliate stigma experienced by family caregivers of people living with dementia.^19^ Although based on testing precise psychometric properties, this scale is limited to the internalized stigma in family caregivers. To understand the stigma process experienced by caregivers, a broader approach is needed that captures their perceptions of stigma from society, reactions to it, and its internalization. This is important for identifying the relationship between perceptions of stigma and its internalization and for exploring the determinants of perceiving being stigmatized by others while resisting it and protecting one’s values and rights.

Bhatt et al. adapted the Family Stigma Instrument (FAMSI), a stigma assessment tool that incorporates these stigma processes, for family caregivers of people living with dementia.^21^ The FAMSI was originally designed for family caregivers of people with intellectual and developmental disabilities.^22,23^ The FAMSI is a multidimensional measure of family stigma, including stigma by association and affiliate stigma.^21^ Additionally, the FAMSI is distinctive in including the positive aspects of caregiving, i.e., beneficial or meaningful experiences related to caregiving, such as personal growth or strengthened relationships.^24,25^ Caregiving is widely accepted to bring about positive reactions and experiences as well as negative ones.^24,25^ The FAMSI is based on both stigma theory and positive psychology caregiving approaches, facilitating understanding of the positive and negative aspects of caregiver influences. This tool may therefore help to understand the complexities of stigma among family caregivers of people living with dementia and the potential role of positive caregiving experiences as its resilience. However, its applicability in Japan remains unclear. In Japan, despite policy emphasis on an inclusive society for people living with dementia and their families, few established tools assess family stigma.

Therefore, this study examined the validity and reliability of the Japanese version of the FAMSI for caregivers of people living with dementia (J-FAMSI-dementia). We translated the FAMSI for dementia into Japanese using a standard translation process and evaluated its suitability in a Japanese sample.

## METHODS

### Study participants

This study included Japanese informal caregivers aged 18–79 years who provided unpaid care for family members living with dementia. Data were collected through an internet-based questionnaire administered via a large survey agency, Macromill Inc. (Tokyo, Japan). We screened 28,313 panelists, of whom 1,669 individuals who engaged in caregiving for family members aged ≥65 years at home were recruited, based on the question: “Are you currently engaged in care for a family member? It includes assisting with caregiving, not only as a primary caregiver.” Caregivers were defined as unpaid caregivers at home; those caregiving for institutionalized persons were excluded. Among them, 899 individuals who responded to the questionnaire were included by age– and sex-stratified sampling. To ensure data quality, 155 respondents who completed the questionnaire in an extremely short time or provided incomplete responses were excluded.^26^ Furthermore, we identified informal caregivers of people living with dementia using the following question: “Does the person receiving care have symptoms suggestive of dementia?” We included the participants who answered either “Yes (diagnosed)” or “Yes (symptoms present but not formally diagnosed).” Finally, 372 informal caregivers were analyzed.

Of the participants, a randomly selected 30% subsample was asked to complete a retest two weeks later using the same questionnaire format. The retest was to assess the stability of the measure over two weeks, which is sufficient time to balance recall bias and unwanted changes.^27^ Ultimately, 103 individuals completed both surveys.

This study was reviewed and approved by the Research Ethics Committee of Hamamatsu University School of Medicine (25-142). Web-based informed consent was obtained from all participants before the survey. This study was conducted in accordance with the principles of the Declaration of Helsinki.

### Family Stigma Instrument

Participants completed the Japanese version of the FAMSI for dementia (J-FAMSI-dementia). The FAMSI for dementia, originally developed in the United Kingdom,^21^ is a 26-item instrument comprising two stigma-related subscales of stigma by association (eight items) and affiliate stigma (perceived, affective, and behavioral subscales with four items each), and a third subscale related to positive aspects of caregiving (six items). Stigma by association refers to caregivers’ recognition of others’ negative evaluations of stigmatized individuals living with dementia and their families (e.g., “Some people might feel embarrassed about associating with people living with dementia”). Affiliate stigma represents the internalization of stigma by caregivers, such that they experience its negative evaluation firsthand (perceived: e.g., “I am excluded from activities when other people find out about their dementia”), experience negative emotions related to their role as caregivers (affective: e.g., “I feel embarrassed about my family member with dementia”), and engagement in negative behaviors, such as avoiding social interactions or being seen as related to the person living with dementia (behavioral: e.g., “I avoid introducing my friends to my family member living with dementia”). Positive aspects of caregiving capture caregivers’ positive attitudes and gains from caregiving (e.g., “Caring for my family member living with dementia has enabled me to develop a more positive attitude toward life”). All items use a 5-point Likert scale ranging from “strongly disagree” (1) to “strongly agree” (5), with a midpoint of “neither agree nor disagree” (3). The original validation of the FAMSI for dementia reported good internal consistency (Cronbach’s alpha = 0.720–0.917) and test-retest reliability (intraclass correlation coefficients [ICCs] = 0.728–0.832).^21^

We translated the original version into Japanese using forward and backward translations. First, two independent researchers in social welfare and health care translated the scale into Japanese. The translations were then reconciled during a joint meeting with another public health researcher, merging differences through consultation. The prepared Japanese version was subsequently back-translated by a third person whose native language was English, and any discrepancies with the original version were resolved through additional forward translation. Finally, the back-translation with the revised Japanese version was reviewed by the original authors to complete the J-FAMSI-dementia.

### Dementia attitude

Dementia attitude was assessed using the Four-Item Attitudes toward People Living with Dementia Scale (APDS4), including items on helping behaviors, sharing feelings, conversations, and interactions with people living with dementia.^28^ The APDS4 uses a 4-point Likert scale, with total scores ranging from 4 to 16; higher scores indicate more positive attitudes toward people living with dementia. The APSDS4 has shown adequate concurrent validity, internal consistency, and test-retest reliability.^28^

### Subjective caregiver burden

Participants completed the Japanese version of the Zarit Burden Interview-8 item scale (J-ZBI_8) to evaluate the perceived burden experienced by caregivers.^29^ Scores range from 0 to 32, with higher scores indicating greater burden. The J-ZBI_8 has demonstrated high concurrent and construct validity and good internal consistency.^29^

### Depressive symptoms

Depressive symptoms were assessed using the Japanese version of the Patient Health Questionnaire-9 (J-PHQ-9) for screening depressive mood, anxiety, and other psychiatric disorders.^30^ Scores range from 0 to 27, with higher scores indicating more severe depressive symptoms. The J-PHQ-9 has shown good sensitivity and specificity for major depression.^30^

### Socio-demographic information

We collected information on age, gender, marital status, educational attainment, subjective economic status, employment status, relationship with the person living with dementia, caregiving roles, and duration of caregiving. Care recipients’ age, gender, care-needs level, type of dementia, and duration of dementia were also collected from caregivers’ reports. In the Japanese public long-term care insurance system, certified individuals are classified into seven levels regarding their care need, with the higher levels indicating greater disabilities (Appendix 1).^31^

### Statistical analysis

First, the response distributions for the 26 J-FAMSI-dementia items were calculated, including the endorsement percentage of “strongly agree” or “agree.” Second, confirmatory factor analysis (CFA) tested the model fit. We examined whether the five-factor structure aligned with the original version. Good fit (and acceptable fit) criteria were in accordance with a previous report^32^: goodness-of-fit index (GFI) >0.95 (0.90); adjusted GFI (AGFI) >0.95 (0.90); root mean square residual (RMR) <0.05 (0.08); standardized root mean residual (SRMS) <0.05 (0.08); comparative fit index (CFI) >0.95 (0.90); Tucker–Lewis index (TLI) >0.95 (0.90); root mean square error of approximation (RMSEA) <0.05 (0.08). Third, Cronbach’s alpha was calculated to assess the internal consistency reliability. Fourth, to evaluate construct validity, we conducted correlation analyses between each J-FAMSI-dementia subscale score and dementia attitude, subjective caregiver burden, and depressive symptoms. We hypothesized that stigma by association and affiliate stigma would be negatively correlated with dementia attitude and positively correlated with caregiver burden and depressive symptoms, whereas positive aspects of caregiving would show the opposite pattern. Finally, to assess the test-retest reliability, we calculated ICCs for subscale scores across two-time responses.

The significance level was set at <0.05. All statistical analyses were conducted using R version 4.3.3 for Windows (R Foundation for Statistical Computing, Vienna, Austria).

## Results

Data from 372 caregivers were analyzed. Participant characteristics are shown in Appendix 2. The mean age was 52.6 years (standard deviation [SD] = 12.2), and 50.0% were women. Among them, 2.2% were spouses/partners, 72.0% were children, and 12.1% were children-in-law, 51.3% were primary caregivers, and 42.2% had been engaged in caregiving for over three years. The care recipients’ mean age was 84.7 years (SD = 6.7), and 68.7% were women. Of the recipients, 88.7% were certified for long-term care, and 32.3% were at care level 3 or higher. Alzheimer’s disease accounted for 43.3%, and 32.8% had lived with dementia for at least three years.

Table 1 presents the endorsement ratings for the J-FAMSI-dementia. The items’ correlation matrix is shown in Appendix 3. All items were approximately normally distributed, with mean scores ranging from 2.4 to 3.4. No clear ceiling or floor effects were observed, and no item showed markedly uneven responses. In the stigma by association subscale, one-third to one-half of caregivers agreed with the statements; particularly, item 2 (feeling uncomfortable about going to the house of the family of someone living with dementia) and item 7 (refusal to hear about the problems of the family of someone living with dementia) were endorsed by 49.7% and 50.2%, respectively. Agreement with the affiliate stigma items ranged from 15.6% to 41.4%, and the perceived and behavioral subscales showed relatively low endorsement, around 20%, whereas the affective subscale items, such as feeling distress about being associated with a family member living with dementia (item 14), were endorsed somewhat more often (17.0% to 41.4%). For positive aspects of caregiving, endorsement ranged from 21.5% to 52.2%, especially for item 22 (caring has made me feel needed: 50.2%) and item 26 (caring has strengthened some of my relationships with family/friends: 40.4%).

**Table 1.** Endorsement ratings of the J-FAMSI-dementia.

| Item wording |  | Mean (SD) | Endorsement of each response option, n (%) |  |  |  |  |
| --- | --- | --- | --- | --- | --- | --- | --- |
|  |  |  | 1 = Strongly disagree | 2 = Somewhat disagree | 3 = Neither disagree/agree | 4 = Somewhat agree | 5 = Strongly agree |
| **Stigma by association** |  |  |  |  |  |  |  |
| 1 | Some people might feel embarrassed about associating with family members of people living with dementia. | 2.9 (1.1) | 31 ( 8.3) | 127 (34.1) | 86 (23.1) | 111 (29.8) | 17 ( 4.6) |
| 2 | Some people might feel uncomfortable about going to the house of the family of someone living with dementia. | 3.2 (1.0) | 21 ( 5.6) | 72 (19.4) | 98 (26.3) | 157 (42.2) | 24 ( 6.5) |
| 3 | Some people might treat the family of someone living with dementia more negatively. | 3.2 (1.0) | 23 ( 6.2) | 84 (22.6) | 104 (28.0) | 136 (36.6) | 25 ( 6.7) |
| 4 | Some people might think that the family has done something wrong because of the person living with dementia. | 2.9 (1.0) | 32 ( 8.6) | 116 (31.2) | 112 (30.1) | 94 (25.3) | 18 ( 4.8) |
| 5 | Some people might behave negatively towards the family when they are with the person living with dementia in public. | 3.1 (1.0) | 14 ( 3.8) | 100 (26.9) | 105 (28.2) | 130 (34.9) | 23 ( 6.2) |
| 6 | Some people might avoid making friends with the family of someone with dementia. | 3.0 (1.0) | 18 ( 4.8) | 118 (31.7) | 110 (29.6) | 109 (29.3) | 17 ( 4.6) |
| 7 | Some people might not want to hear about any of the problems of the family of someone living with dementia. | 3.3 (1.0) | 10 ( 2.7) | 67 (18.0) | 112 (30.1) | 155 (41.7) | 28 ( 7.5) |
| 8 | Some people might not invite the family of someone living with dementia to social events. | 3.2 (1.0) | 15 ( 4.0) | 92 (24.7) | 110 (29.6) | 123 (33.1) | 32 ( 8.6) |
| **Perceived affiliate stigma** |  |  |  |  |  |  |  |
| 9 | I am treated differently by some people when I am with my family member living with dementia. | 2.7 (1.0) | 33 ( 8.9) | 135 (36.3) | 121 (32.5) | 71 (19.1) | 12 ( 3.2) |
| 10 | I am excluded from activities when other people find out about their dementia. | 2.5 (1.0) | 56 (15.1) | 140 (37.6) | 118 (31.7) | 46 (12.4) | 12 ( 3.2) |
| 11 | I am aware of how some people look at me when I am out with my family member living with dementia. | 2.7 (1.1) | 44 (11.8) | 142 (38.2) | 97 (26.1) | 69 (18.5) | 20 ( 5.4) |
| 12 | I am treated differently by some people because of my family member living with dementia. | 2.7 (1.0) | 32 ( 8.6) | 139 (37.4) | 124 (33.3) | 65 (17.5) | 12 ( 3.2) |
| **Affective affiliate stigma** |  |  |  |  |  |  |  |
| 13 | I feel embarrassed about my family member living with dementia. | 2.7 (1.0) | 32 ( 8.6) | 132 (35.5) | 123 (33.1) | 73 (19.6) | 12 ( 3.2) |
| 14 | I feel distressed about being associated with my family living with dementia. | 3.2 (1.0) | 19 ( 5.1) | 85 (22.8) | 114 (30.6) | 123 (33.1) | 31 ( 8.3) |
| 15 | I feel guilty about having my family member living with dementia in the family. | 2.5 (1.0) | 56 (15.1) | 163 (43.8) | 90 (24.2) | 52 (14.0) | 11 ( 3.0) |
| 16 | I feel uncomfortable when I have friends about because of my family member living with dementia. | 2.9 (1.0) | 28 ( 7.5) | 101 (27.2) | 148 (39.8) | 79 (21.2) | 16 ( 4.3) |
| **Behavioral affiliate stigma** |  |  |  |  |  |  |  |
| 17 | I avoid introducing my friends to my family member living with dementia. | 2.7 (1.0) | 41 (11.0) | 122 (32.8) | 131 (35.2) | 59 (15.9) | 19 ( 5.1) |
| 18 | I avoid telling people that I am related to my family member living with dementia. | 2.6 (1.0) | 46 (12.4) | 149 (40.1) | 108 (29.0) | 54 (14.5) | 15 ( 4.0) |
| 19 | I avoid making new friends because of my family member living with dementia. | 2.5 (1.0) | 54 (14.5) | 144 (38.7) | 106 (28.5) | 56 (15.1) | 12 ( 3.2) |
| 20 | I avoid being seen with my family member living with dementia. | 2.4 (1.0) | 65 (17.5) | 153 (41.1) | 96 (25.8) | 45 (12.1) | 13 ( 3.5) |
| **Positive aspects of caregiving** |  |  |  |  |  |  |  |
| 21 | Caring for my family member living with dementia has enabled me to develop a more positive attitude toward life. | 3.0 (1.0) | 27 ( 7.3) | 87 (23.4) | 151 (40.6) | 91 (24.5) | 16 ( 4.3) |
| 22 | Caring for my family member living with dementia has made me feel needed. | 3.4 (1.0) | 19 ( 5.1) | 56 (15.1) | 110 (29.6) | 150 (40.3) | 37 ( 9.9) |
| 23 | Caring for my family member living with dementia has strengthened my spirituality and faith. | 2.8 (1.1) | 43 (11.6) | 109 (29.3) | 131 (35.2) | 69 (18.5) | 20 ( 5.4) |
| 24 | Caring for my family member living with dementia has allowed me to form friendships with others in a similar situation. | 3.0 (1.0) | 31 ( 8.3) | 80 (21.5) | 139 (37.4) | 100 (26.9) | 22 ( 5.9) |
| 25 | Caring for my family member living with dementia has made me feel that I make a positive contribution to society. | 2.7 (1.0) | 42 (11.3) | 108 (29.0) | 142 (38.2) | 64 (17.2) | 16 ( 4.3) |
| 26 | Caring for my family member living with dementia has strengthened some of my relationships with family/friends. | 3.1 (1.1) | 34 ( 9.1) | 71 (19.1) | 117 (31.5) | 123 (33.1) | 27 ( 7.3) |

J-FAMSI-dementia, Japanese version of the Family Stigma Instrument for dementia; SD, standard deviation.

Figure 1 illustrates the CFA of the J-FAMSI-dementia. The five-factor structural model as presented in the original version showed acceptable model fit (chi-square = 709.700, *P* <0.001; GFI = 0.867; AGFI = 0.838; RMR = 0.058; SRMR = 0.057; CFI = 0.919; TLI = 0.908; RMSEA = 0.063). Each factor indicated high internal consistency (Cronbach’s alpha: stigma by association = 0.878; perceived affiliate stigma = 0.855; affective affiliate stigma = 0.814; behavioral affiliate stigma = 0.886; positive aspects of caregiving = 0.863; affiliate stigma subtotal = 0.930).

**Figure 1.**
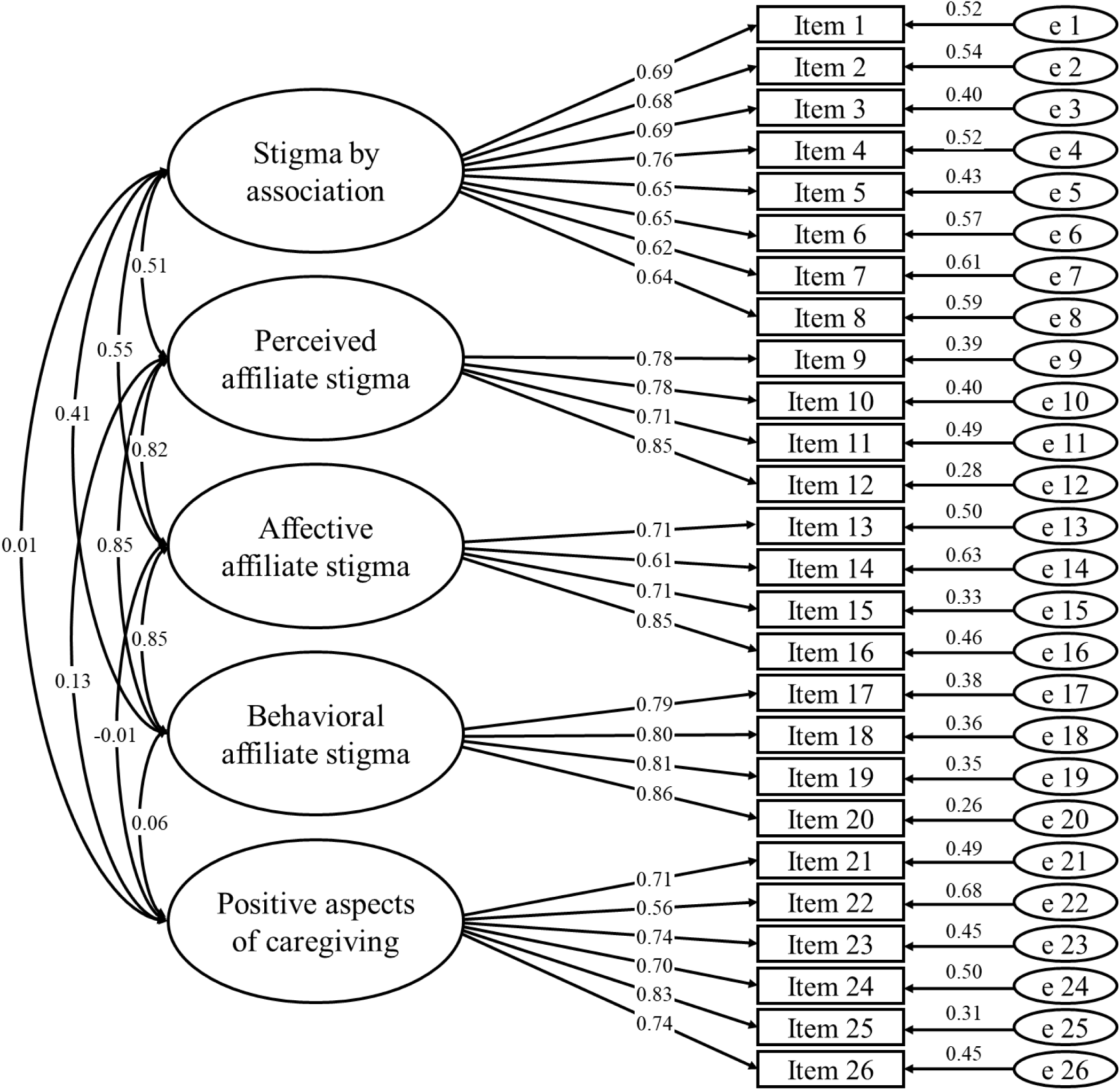
Confirmatory factor analytical model of the Japanese version of the Family Stigma Instrument for dementia (J-FAMSI-dementia), using maximum likelihood estimates. Numbers on the lines between factors indicate covariance between factors, and numbers on arrows from factors to scale items indicate standardized factor loading estimates. Chi-square = 709.700, *P* <0.001; goodness-of-fit index (GFI) = 0.867; adjusted GFI (AGFI) = 0.838; root mean square residual (RMR) = 0.058; standardized root mean residual (SRMR) = 0.057; comparative fit index (CFI) = 0.919; Tucker-Lewis index (TLI) = 0.908; root mean square error of approximation (RMSEA) = 0.063.

Table 2 indicates the correlations among the J-FAMSI-dementia subscales. The four stigma-related factors were positively intercorrelated. Stigma by association was moderately correlated with affiliate stigma factors (r = 0.382 to 0.489), and affiliate stigma factors were moderately to strongly correlated with each other (r = 0.689 to 0.757). Contrary, positive aspects of caregiving showed little correlation with stigma factors (r = –0.058 to 0.089).

**Table 2.** Correlations between the J-FAMSI-dementia subscales.

|  | Correlation coefficients |  |  |  |
| --- | --- | --- | --- | --- |
|  | Stigma by association | Perceived affiliate stigma | Affective affiliate stigma | Behavioral affiliate stigma |
| Stigma by association | – |  |  |  |
| Perceived affiliate stigma | 0.455*** | – |  |  |
| Affective affiliate stigma | 0.489*** | 0.689*** | – |  |
| Behavioral affiliate stigma | 0.382*** | 0.757*** | 0.702*** | – |
| Positive aspects of caregiving | 0.013 | 0.089 | -0.058 | 0.035 |

\*\*\*Pearson's correlation significant at $P < 0.001$ .
J-FAMSI-dementia, Japanese version of the Family Stigma Instrument for dementia.

Table 3 presents correlations between the J-FAMSI-dementia and external criterion variables. Stigma by association and affiliate stigma factors were weakly negatively correlated with dementia attitude (r = –0.193 to –0.389), whereas positive aspects of caregiving were positively correlated (r = 0.315). For subjective caregiver burden, stigma by association showed a weak positive correlation (r = 0.285), and affiliate stigma factors showed moderate positive correlations (r = 0.386 to 0.496); positive aspects of caregiving showed weak negative correlations (r = –0.165). Regarding depressive symptoms, stigma by association and affiliate stigma factors showed weak positive correlations (r = 0.196 to 0.377), whereas positive aspects of caregiving showed weakly negative correlations (r = –0.203).

**Table 3.** Correlations between the J-FAMSI-dementia and external criterion variables.

|  | Correlation coefficients |  |  |
| --- | --- | --- | --- |
|  | Dementia attitude | Subjective caregiver burden | Depressive symptoms |
| Stigma by association | -0.193*** | 0.285*** | 0.196*** |
| Perceived affiliate stigma | -0.239*** | 0.403*** | 0.354*** |
| Affective affiliate stigma | -0.361*** | 0.496*** | 0.320*** |
| Behavioral affiliate stigma | -0.389*** | 0.386*** | 0.344*** |
| Affiliate stigma (subtotal) | -0.366*** | 0.473*** | 0.377*** |
| Positive aspects of caregiving | 0.315*** | -0.165** | -0.203*** |

\*\*\*Pearson's correlation significant at $P < 0.001$ .
J-FAMSI-dementia, Japanese version of the Family Stigma Instrument-dementia.

Table 4 indicates the test-retest scores over a two-week interval. The ICCs for affective and behavioral affiliate stigma, positive aspects of caregiving, and the subtotal of affiliate stigma were 0.70 to 0.77, indicating good reliability. Stigma by association and perceived affiliate stigma showed moderate reliability (ICCs = 0.61 and 0.69, respectively).

**Table 4.** Scores of the J-FAMSI-dementia on two occasions (n = 103)

|  | Score range | First administration |  | Second administration |  | ICC (95% CI) | *P*-value |
| --- | --- | --- | --- | --- | --- | --- | --- |
|  |  | Mean (SD) | Observed range | Mean (SD) | Observed range |  |  |
| Stigma by association | 8–40 | 25.6 (5.4) | 12–40 | 25.8 (5.8) | 9–37 | 0.61 (0.48–0.72) | < 0.001 |
| Perceived affiliate stigma | 4–20 | 10.6 (3.3) | 4–20 | 11.2 (3.5) | 4–20 | 0.69 (0.57–0.78) | < 0.001 |
| Affective affiliate stigma | 4–20 | 11.4 (3.0) | 4–20 | 11.5 (3.2) | 4–19 | 0.75 (0.65–0.82) | < 0.001 |
| Behavioral affiliate stigma | 4–20 | 10.2 (3.6) | 4–20 | 10.7 (3.7) | 4–20 | 0.77 (0.67–0.84) | < 0.001 |
| Affiliate stigma (total) | 12–60 | 32.1 (9.1) | 13–60 | 33.2 (9.7) | 13–58 | 0.75 (0.65–0.82) | < 0.001 |
| Positive aspects of caregiving | 6–30 | 18.1 (4.7) | 6–28 | 18.0 (4.9) | 6–28 | 0.70 (0.59–0.79) | < 0.001 |

CI, confidence interval; ICC, intraclass correlation coefficient; J-FAMSI-dementia, Japanese version of the Family Stigma Instrument-dementia; SD, standard deviation

## Discussion

This study developed the Japanese version of the FAMSI-dementia, a multidimensional stigma scale for family caregivers of people living with dementia. The J-FAMSI-dementia indicated acceptable factor validity, reasonable concurrent validity, and good reliability. Establishing the J-FAMSI-dementia may help identify stigma among family caregivers of people living with dementia.

The J-FAMSI-dementia showed a factorial structure similar to the original version,^21^ with acceptable validity and good reliability. These psychometric properties support the suitability of this scale for family caregivers of people living with dementia in the context of Japanese society. Previous scales of family stigma have focused only on the family’s negative experiences and affiliate stigma,^19,20^ which are insufficient to deepen understanding of the full scope of family caregiving experiences related to dementia stigma. The FAMSI-dementia covers both aspects of stigma by association as directed by society and affiliate stigma as its internalization, thereby capturing the complex psychological processes of family caregivers of people living with dementia. Additionally, the positive framework of caregiving contained in the FAMSI-dementia may help to comprehend the balance between negative and positive experiences of family caregivers and investigate strategies to resist stigma from society and stigmatized beliefs and behaviors. Although several caregiver stress models are widely used,^33,34^ there remains a lack of perspective on the stigma experienced by family caregivers. Assessing family stigma deepens understanding of psychological distress in dementia caregiving and contributes to developing a caregiver support model incorporating stigma.

Each J-FAMSI-dementia subscale correlated with external indicators, as hypothesized. The stigma-related subscales were inversely correlated with positive attitudes toward dementia and positively correlated with caregiver burden and depressive symptoms, whereas positive aspects of caregiving showed the opposite pattern. These partially support the criterion-related validity of the scale. Although the correlations were weak to moderate, this suggests a degree of discrimination in that the stigma dimensions are not entirely explained by attitudes, burden, or mental health.

Correlations within affiliate stigma subscales were stronger than those between stigma by association and affiliate stigma. This is plausible in reflecting the cohesiveness of internalized stigmas. According to the hypothesis of the cognitive-behavioral model,^35,36^ affiliate stigma includes three aspects of perceived, affective, and behavioral. After becoming aware of stigma from society, caregivers undergo internalization processes in which they experience this firsthand (perceived), experience negative emotions related to their caregiving role (affective), and engage in stigma-related behaviors, such as avoiding social interactions and hiding the presence of family members living with dementia. These internalizations are assumed to arise from stigma by association, namely recognition of prejudice and discrimination directed at families of people living with dementia by society. Our results support that the scale captures distinct yet related aspects of associative and affiliate stigma.

The positive aspects of caregiving subscale did not correlate with the other stigma subscales. This is partly consistent with research in the field of intellectual disabilities showing that positive aspects of caregiving are only weakly related to or little explained by stigma.^22^ Still, while one-quarter to one-half of participants experienced stigma, a similar proportion endorsed the positive aspects of caregiving. This suggests that stigma and positive caregiving feelings were experienced at different levels, not along the same axis. Although understanding the role of the positive aspects of caregiving in relation to stigma requires further investigations, fostering them may represent a positive coping strategy against internalized stigma.

Nearly half of the caregivers affirmed stigma by association, suggesting their awareness of stigmatizing views and treatment from others and society due to their relationship with the person living with dementia. Particularly, caregivers often endorsed the items about others refusing to hear about their problems or feeling uncomfortable going to their homes. These responses suggest that the general public may distance themselves psychosocially from people who have family members living with dementia and that caregivers perceive this sensitively. In contrast, caregivers affirmed affiliate stigma somewhat less often. This pattern suggests a complex relationship in which societal stigma does not necessarily lead to internalization. Notably, the affective dimension of affiliate stigma was affirmed somewhat more often than the perceived and behavioral dimensions, which is a different pattern from previous research in the United Kingdom.^21,22^ In Japanese society, which places importance on public appearance,^37^ having a family member with dementia might cause negative psychological consequences for caregivers, such as shame and distress, even when caregivers do not strongly perceive direct affiliate stigma. This complicated psychosocial suffering experienced by caregivers may show differential trends from that reported in Western countries and may reflect broader Asian cultural contexts, including Japan, as a collective society with latent familism beliefs.^38^ These findings highlight the need to further investigate these cultural differences.

This study has several limitations. First, the participants were limited to informal caregivers of family members aged ≥65 years living with dementia. It is unclear whether this scale is adaptable to caregivers of people with early-onset dementia. Second, information on the care recipients’ dementia was based on caregivers’ reports rather than clinical diagnosis, which may have caused misclassification. Still, because some caregivers support people with possible but undiagnosed dementia in the community,^39^ this study was also able to cover this relevant population. Third, the participant characteristics recruited through an internet research agency were not necessarily representative of the general caregiver population. Particularly, spousal caregivers were underrepresented. This may differ from the situation in Japan, where older couples often care for each other.^40^ It needs to be verified in population-based samples.

## CONCLUSION

This study developed the J-FAMSI-dementia, a Japanese stigma scale for family caregivers of people living with dementia, and demonstrated its validity and reliability. This instrument may help deepen understanding of the stigma experienced in dementia care and facilitate efforts to reduce stigma and strengthen caregiver support.

## Acknowledgements

We wish to express our sincere gratitude to Prof. Katrina Scior, University College London, for the contribution to this study. We also thank Macromill, Inc., all participants in this study, and Crimson Interactive Pvt. Ltd. (Ulatus; www.ulatus.jp) for their assistance in manuscript translation and editing.

## Author contributions

TN conceptualized and designed the study, participated in data collection, analyzed the data, and drafted and revised the manuscript. JB, AK, and RW supported designing the study and reviewed and critically revised the manuscript. All authors approved the submission of the final manuscript.

## Data availability statement

The data that support the findings of this study are available on request from the corresponding author. The data is not publicly available due to privacy or ethical restrictions.

