## Appendix for "Japanese version of the Family Stigma Instrument for informal caregivers of people with dementia"

**Appendix 1. Definitions of the care-needs levels used in the Japanese public long-term care insurance system**

| Level | Total daily estimated care minutes | ADL conditions |
| --- | --- | --- |
| Support level 1 | 25–32 | Long-term care is needed for some aspects of daily living, but proper care can improve or maintain ADL |
| Support level 2 | 32–50 |  |
| Care level 1 | 32–50 | Unstable in rising and gait; partial support needed in toileting, bathing, etc. |
| Care level 2 | 50–70 | Difficulty in rising and gaiting; partial or complete support needed in toileting, bathing, etc. |
| Care level 3 | 70–90 | Impossible to rise and no gait. Complete support needed in toileting, bathing, dressing, and all other basic ADL |
| Care level 4 | 90–110 | Severe decline in ADL capacity; complete support needed in toileting, bathing, dressing, and all other basic ADL |
| Care level 5 | ≥ 110 | Complete support needed in all ADL; difficulty with communication |

ADL, activities of daily living.

**Appendix 2. The characteristics of the study participants**

|  |  | Overall (n = 372) |
| --- | --- | --- |
|  |  | n (%) |
| **Caregivers** |  |  |
| Age (years), mean (SD) |  | 52.6 (12.2); range 18–79 |
| Gender | Men | 186 ( 50.0) |
|  | Women | 186 ( 50.0) |
| Marital status | Married | 195 ( 52.4) |
|  | Separated/divorced | 49 ( 13.2) |
|  | Never married | 125 ( 33.6) |
|  | Others | 3 ( 0.8) |
| Educational attainment (years) | < 10 | 102 ( 27.4) |
|  | 10–12 | 86 ( 23.1) |
|  | ≥ 13 | 184 ( 49.5) |
| Subjective economic status | Severe | 76 ( 20.4) |
|  | Somewhat severe | 102 ( 27.4) |
|  | Normal | 147 ( 39.5) |
|  | Somewhat afford | 36 ( 9.7) |
|  | Afford | 11 ( 3.0) |
| Employment status | Regular employed | 150 ( 40.3) |
|  | Self-employed | 51 ( 13.7) |
|  | Not regular employed | 75 ( 20.2) |
|  | Not employed | 85 ( 22.8) |
|  | Others | 11 ( 3.0) |
| Relationship to a person living with dementia | Spouse/partner | 8 ( 2.2) |
|  | Child | 268 ( 72.0) |
|  | Child-in-law | 45 ( 12.1) |
|  | Others | 51 ( 13.7) |
| Caregiving roles | Secondary | 181 ( 48.7) |
|  | Primary | 191 ( 51.3) |
| Caregiving durations | Under 6 months | 23 ( 6.2) |
|  | 6 months to 1 years | 56 ( 15.1) |
|  | 1 to 3 years | 136 ( 36.6) |
|  | 3 to 5 years | 89 ( 23.9) |
|  | 5 to 10 years | 45 ( 12.1) |
|  | Over 10 years | 23 ( 6.2) |
| **Care recipients** |  |  |
| Age (years), mean (SD) |  | 84.7 (6.7); range 65–100 |
| Gender | Male | 118 ( 31.7) |
|  | Female | 254 ( 68.3) |
| Care-needs level | Not certified | 26 ( 7.0) |
|  | Support level 1 | 26 ( 7.0) |
|  | Support level 2 | 24 ( 6.5) |
|  | Care level 1 | 79 ( 21.2) |
|  | Care level 2 | 81 ( 21.8) |
|  | Care level 3 | 71 ( 19.1) |
|  | Care level 4 | 32 ( 8.6) |
|  | Care level 5 | 17 ( 4.6) |
|  | Unknown | 16 ( 4.3) |
| Type of dementia | Alzheimer’s disease | 161 ( 43.3) |
|  | Vascular dementia | 22 ( 5.9) |
|  | Lewy body | 17 ( 4.6) |
|  | Frontotemporal dementia | 14 ( 3.8) |
|  | Others | 17 ( 4.6) |
|  | Mixed | 23 ( 6.2) |
|  | Unknown | 118 ( 31.7) |
| Duration of dementia | Under 6 months | 41 ( 11.0) |
|  | 6 months to 1 years | 68 ( 18.3) |
|  | 1 to 3 years | 142 ( 38.2) |
|  | 3 to 5 years | 62 ( 16.7) |
|  | Over 5 years | 59 ( 15.9) |

SD, standard deviation.

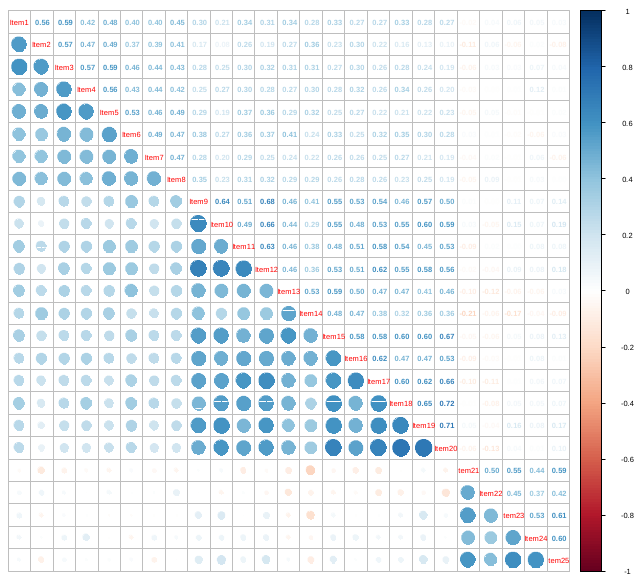

**Appendix 3. Correlation matrix of the Japanese version of the Family Stigma Instrument for dementia (J-FAMSI-dementia).** Circles in the lower triangle and numeric values in the upper triangle represent Pearson correlation coefficients between all items. Circle size and color intensity indicate the magnitude of correlations (blue = positive, red = negative). Diagonal elements are omitted.
